# Ischemic Stroke Risk in Patients on Direct Oral Anticoagulants with Levetiracetam: A Pharmacovigilance Study

**DOI:** 10.1101/2023.08.21.23294397

**Authors:** Mohammed Abou Kaoud, Ran Nissan, Amitai Segev, Avi Sabbag, David Orion, Elad Maor

## Abstract

**Background:** Levetiracetam is widely used in post stroke epilepsy. However, it is suspected to possess P-glycoprotein induction properties and therefore a potential significant interaction with DOACs. Our aim was to search for ischemic stroke signals with levetiracetam and the DOACs.

**Methods:** In this retrospective, pharmacovigilance study, we used the Food and Drug Administration adverse event reporting system to identify ischemic stroke events associated with DOACs and concomitant use of levetiracetam. We evaluated disproportionate reporting by the reporting odds ratio adjusted to age and sex (adj.ROR) and the lower bound of the shrinkage 95% confidence interval (Ω_025_ > 0 is deemed significant for an interaction).

**Results:** We identified 1,841 (1.5%), 3,731 (5.3%), 338 (4.9%), and 1,723 (1.3%), ischemic stroke reports with apixaban, dabigatran, edoxaban, and rivaroxaban respectively. When heparin/enoxaparin was used as the comparator the adjusted ROR of the interaction effect was 3.57 (95%CI, 2.81–4.58) between DOACs and levetiracetam. The shrinkage analysis detected an interaction between each of the DOACs and levetiracetam resulting in higher reports of ischemic stroke with the combination compared to each drug alone. The logistic model and shrinkage analysis failed to detect an interaction when queried for hemorrhagic stroke.

**Conclusions:** We show a strong signal for the levetiracetam interaction with apixaban, dabigatran, edoxaban, and rivaroxaban leading to a 3-5 folds increased reporting risk of ischemic stroke. Our findings suggest the need for pharmacodynamic monitoring, while concomitantly prescribing levetiracetam with the DOACs.

## INTRODUCTION

The introduction of direct oral anticoagulants (DOACs) has significantly altered the management of atrial fibrillation (AF) due to their improved safety profile and ease of use compared with warfarin. Contemporary guidelines prefer DOACs over warfarin for stroke prevention among patients with AF, excluding patients with mechanical valve replacement or significant mitral stenosis (1). While there is no preference for one specific DOAC over another, in the clinic DOACs are personalized according to patient age, gastrointestinal bleeding risk, chronic kidney disease, and drug interactions (2).

Since stroke is one of the most common structural etiologies of epilepsy, the concomitant use of DOACs and an anticonvulsant is not uncommon (3). Post-stroke seizures account for 11% of all epilepsy, 22% of all cases of status epilepticus, and 55% of newly diagnosed seizures amongst older people (4, 5, 6). Since AF is a common cause of ischemic stroke, the rate of AF-stroke-epilepsy triplet, and thus DOAC-anticonvulsant treatment, is not uncommon among older adults (7).

To date, the combination of DOAC and some anticonvulsants remains controversial. The European Heart Rhythm Association (EHRA) 2018 guide did not support using anticonvulsants (carbamazepine, levetiracetam, phenobarbital, phenytoin, topiramate, and valproic acid) in patients concurrently taking DOACs (8). The 2021 EHRA guide states that after inquiry with the drug manufacturer there is unfortunately no study which reliably investigated the effect of levetiracetam on DOAC plasma levels and clinical events in a sufficiently large ‘real world’ cohort of concomitantly treated patients (33). However, in practice, levetiracetam is widely used for post-stroke epilepsy due to its favorable safety profile and fewer drug interactions.

Therefore, our study aim was to investigate reports to the Federal Drug Agency (FDA) regarding ischemic and non-ischemic stroke with and without levetiracetam combination in AF patients treated with DOACs.

## METHODS

An observational, retrospective pharmacovigilance study was carried out using the FAERS database, a global repository of voluntary reports by healthcare professionals and consumers, and mandatory reports from manufacturers (9). The database was screened for reports containing the following terms in their brand or generic names: ‘rivaroxaban,’ ‘apixaban,’ ‘dabigatran,’ ‘dabigatran etexilate mesylate,’ dabigatran etexilate,’ ‘edoxaban,’ ‘edoxaban mesylate,’ ‘edoxaban tosylate,’ ‘enoxaparin,’ ‘enoxaparin sodium,’ ‘heparin,’ ‘heparin sodium,’ ‘heparin calcium,’ ‘levetiracetam,’ ‘carbamazepine’ and ‘omeprazole.’ Omeprazole and carbamazepine are, respectively, negative and positive controls in this study.

The study included patients who were reported as the primary suspects for a given adverse event (AE) when novel anticoagulants accessed the market (2012–2023). In case multiple reports of the same event were detected, only the latest case version of every event was retained, as recommended by the FDA. We further applied a population-linkage program to detect suspected duplicate reports of the same drug-event pair with different case numbers by screening for identical values in six key fields: age, sex, event date, country of occurrence, concomitant medications, and the same reasons for use. These suspected duplicate reports were excluded.

The database was then searched for the following adverse events in their Medical Dictionary for Regulatory Activities (MedDRA) preferred terms: ‘ischemic stroke,’ ‘hemorrhagic stroke,’ ‘cerebral hemorrhage,’ ‘intracranial hemorrhage,’ ‘cerebrovascular accident,’ ‘lacunar stroke,’ ‘cerebellar stroke,’ ‘cerebral artery occlusion,’ ‘basal ganglia stroke’, ‘vertebrobasilar stroke’, ‘brain stem stroke’, ‘thrombotic stroke’ and ‘embolic stroke.’ Those adverse events either denote a lack of effectiveness or toxicity. Detailed lists of terms included in our database query are available at http://bioportal.bioontology.org/.

The data was then filtered by DOAC’s reason for use: embolic stroke in AF patients’ treatment and prophylaxis. The dataset was then queried for cases with concomitant anticonvulsants used. Cases involving anticonvulsants were identified by a predefined list of anticonvulsant medications constructed using the FDA National Drug Code (NDC) file (10). Only patients above age 12 and cases with one reported anticonvulsant were included.

### Study End Points

The predefined primary endpoint was any MedDRA-preferred term describing ischemic stroke. Additional information for each report in the database, including demographic information (country, reporter occupation, reporting year, age, and sex), concomitant anticonvulsant uses, and date of AE occurrence and its outcomes, were collected for the analysis. Cases were defined as serious medical events if one or more of the following outcomes were reported: death, life-threatening event, hospitalization, disability, or another serious medical event.

### Statistical Analysis

Descriptive statistics were calculated for patient demographics. Means and Standard deviations were generated for continuous variables. Frequencies and proportions were stated for categorical variables. A validated case-non-case method in drug safety research assessed whether ischemic and non-ischemic strokes were reported more frequently with the DOACs: apixaban, edoxaban, rivaroxaban, or dabigatran, compared with enoxaparin or heparin. We further analyzed each of the DOACs compared to the other three for reported ischemic stroke.

A logistic regression model obtained an adjusted reporting odds ratio (adj. ROR). The regression model included age, sex, and concomitant levetiracetam use. The ROR is a statistical model surrogate to the odds ratio and is an acceptable method to detect signals of adverse events and drug interactions (11, 12). Missing sex data were coded as ‘Not Specified,’ and the median value imputed missing age data in each group of drug recipients. R-squared and adjusted R-squared were used to assess model fitness. The ROR for concomitant levetiracetam was used to measure the strength of the association of the drug interaction.

Additionally, a refined model called the Ω shrinkage measure was used to calculate the observed-to-expected for detecting signals of potential drug-drug interactions (DDIs). Omega Ω is a robust observed-to-expected triplet measure of disproportionate reporting developed by the Uppsala Monitoring Centre (12, 13). When Ω is positive, and two drugs are used together, an increased risk of a specific adverse event occurrence is emphasized over the sum of the individual risks when these same drugs are used separately (13). Thus, Ω_025_ > 0, a positive lower bound of 95% CI, is used as a threshold for detecting the signals of the concomitant use of drug D_1_ and drug D_2_ (12). In other words, it indicates the frequency of reporting specific Drug-Drug-Event triplets in the dataset compared to what is expected based on the relative reporting for each drug alone (see eq. 1).

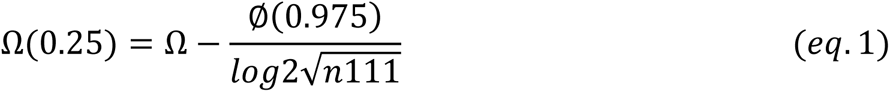

Where ∅(0.975) is the standard normal distribution, and n111 is the number of cases with the Drug-Drug-Event triplet.

The logistic regression analyses were performed using SPSS version 27. Two-sided *P* < .05 indicated significance. Shrinkage calculation steps were inputted in Microsoft Excel version 2022. Noguchi Y. et al. 2019 described the complete shrinkage calculation (12).

### Sensitivity Analysis

First, we examined whether the drug interaction also manifests as hemorrhagic stroke. A model for hemorrhagic stroke comparing each of the DOACs; apixaban, rivaroxaban and dabigatran against the other two was performed. Edoxaban was not included in this analysis since there were no reports of hemorrhagic events with the levetiracetam-edoxaban combination.

A sensitivity analysis for the levetiracetam drug interaction, included two drug combinations: apixaban-carbamazepine and apixaban-omeprazole. The former drug interaction is well established. Carbamazepine is known to induce P-glycoprotein and CYP3A4, thus reducing apixaban effectiveness (FDA and European Medical Agency physician guidance; 14,15). In contrast to carbamazepine, the combination of apixaban and omeprazole is widespread and considered safe without any known or potential drug interaction (16).

We further stratified DOACs by publication date in an additional analysis up to 2020, aiming to mitigate reporting bias. We hypothesized that from 2020 onwards, increased reports regarding DOACs, and ischemic stroke may be expected since the publication by Giustazzi et al. showed decreased effectiveness with concomitant DOAC-anticonvulsant use. Rivaroxaban was selected for further analysis since most patients in the Giustazzi et al. study were treated with rivaroxaban and more case reports were published regarding rivaroxaban (17).

Further, we re-analyzed our results for the period 2015-2023, this is to account for the fact that during 2010-2014 a trial had been ongoing against Boehringer Ingelheim, dabigatran’s manufacturer (18). Thus, to mitigate any additional negative reporting bias, we analyzed our data excluding the years 2012-2014.

### Standard Protocol Approvals, Registrations, and Patient Consents

The FAERS Public Dashboard is a publicly available web-based tool containing mandatory data reports from drug manufacturers and voluntary ADR reports from consumers and healthcare professionals. Hence an informed consent or ethical statement of approval by an ethical standards committee on human experimentation is not required for analysis.

## RESULTS

### Demographics and Patient Characteristics

The FAERS database included 19,609,956 unique safety reports from January 2012 to January 2023. Out of these reports, 125,799 for Apixaban, 69,993 for Dabigatran, 6,965 for Edoxaban, and 136,710 for Rivaroxaban. Among eligible patients in the FAERS database that reported any of the MedDRA preferred terms for ischemic stroke are 1,841 (1.5%), 3,731 (5.3%), 338 (4.9%), and 1,723 (1.3%), treated with apixaban, dabigatran, edoxaban, and rivaroxaban respectively. In contrast, reports that described hemorrhagic stroke included 3,016 (2.2%), 1,346 (1.1%), and 1,256 (1.8%) for rivaroxaban, apixaban, and dabigatran. An additional 383 (6.9%) ischemic stroke cases were identified among enoxaparin/heparin treated patients.

The proportion on man significantly higher in both adverse events. The mean age for apixaban-treated patients was 77±10, significantly older than the other three DOACs. Enoxaparin/heparin patients were younger with a mean age of 72±13 and did not differ by sex from the DOACs. Hemorrhagic stroke resulted in poorer outcomes and more mortality compared to ischemic stroke (Table 1).

**Table 1:** Characteristics and Demographics of Levetiracetam and Direct Anticoagulant Users with Associated Ischemic and non-Ischemic Stroke Adverse Events in Atrial Fibrillation Patients.

| (No. of total AEs) | Rivaroxaban<br>(136,710) | Apixaban<br>(125,799) | Dabigatran<br>(69,993) | Edoxaban<br>(6,965) | P-Value |
| --- | --- | --- | --- | --- | --- |
| Total AERs |  |  |  |  |  |
| Ischemic stroke | 1723 (1.3) | 1841 (1.5) | 3731 (5.3) | 338 (4.9) | - |
| Hemorrhagic stroke | 3016 (2.2) | 1346 (1.1) | 1256 (1.79) | - |  |
| Age | 73.6±13.2 | 76.9±9.9 | 73.7±10.6 | 74.5±11.5 | <0.001,<br>0.94 |
| Sex |  |  |  |  |  |
| Female | 794 (46.1) | 772 (46.5) | 1478 (40.1) | 46 (13.6) | <0.001 |
|  | 1287 (42.7) | 553 (41.1) | 518 (41.2) | - |  |
| Male | 823 (47.8) | 791 (47.7) | 1881 (51.1) | 51 (15.0) |  |
|  | 1641 (54.5) | 737 (54.8) | 683 (54.4) | - |  |
| Not Specified | 106 (6.2) | 97 (5.8) | 325 (8.8) | 241 (71.3) |  |
|  | 88 (2.9) | 28 (4.2) | 55 (4.4) | - |  |
| No. concomitant Drug-AE |  |  |  |  |  |
| Levetiracetam | 34 (2.0) | 32 (1.7) | 33 (0.9) | 4 (1.2) | - |
| Ischemic | 14 (0.5) | 10 (0.7) | 13 (1.0) | N/R |  |
| Levetiracetam |  |  |  |  |  |
| Hemorrhagic |  |  |  |  |  |
| Mortality outcome<sup>b</sup> |  |  |  |  |  |
| Ischemic stroke |  |  |  |  | <0.001 |
| DOAC alone | 305 (11.8) | 111 (6.0) | 451 (7.7) | 23 (6.8) |  |
| DOAC-levetiracetam | 1 (0.04) | 4 (0.2) | 12 (0.2) | 0 (0) |  |
| Hemorrhagic stroke |  |  |  |  |  |
| DOACs alone | 1552 (56.9) | 488 (36.3) | 588 (46.8) | - |  |
| DOAC-levetiracetam | 20 (0.7) | 4 (0.3) | 7 (0.6) | N/R |  |
| Publication date |  |  |  |  |  |
| 2012-2013 | 141 (8.2) | 16 (0.87) | 971 (26.0) | 446 (6.4) | <0.001 |
|  | 0 (0) | 7 (0.5) | 305 (24.3) |  |  |
| 2014-2015 | 355 (20.6) | 201 (10.9) | 749 (20.1) | 1526 (21.9) |  |
|  | 339 (11.2) | 8 (0.6) | 276 (21.0) |  |  |
| 2016-2017 | 571 (33.1) | 408 (22.2) | 582 (15.6) | 1832 (26.3) |  |
|  | 1072 (35.5) | 313 (23.3) | 215 (17.1) |  |  |
| 2018-2019 | 383 (22.2) | 493 (26.8) | 725 (19.4) | 1651 (23.7) |  |
|  | 689 (22.8) | 584 (43.4) | 249 (19.8) |  |  |
| 2020-2021 | 244 (14.2) | 386 (21.0) | 458 (12.2) | 1468 (21.1) |  |
| 2022-2023 | 854 (28.3)<br>29 (1.68)<br>62 (2.1) | 313 (23.2)<br>157 (8.5)<br>120 (8.9) | 115 (9.2)<br>99 (2.7)<br>11 (0.9) | 72 (1.0) |  |

AEs-adverse events. Top figures for ischemic stroke, bottom figures for hemorrhagic stroke. Numbers in brackets are percentages of ischemic stroke events.

Concomitant levetiracetam reports and percentages of the total in each anticoagulant were as follows: 122 (0.1%) in apixaban, 142 (0.2%) in dabigatran, 20 (0.3%) in edoxaban, 168 (0.1%) in rivaroxaban, and 45 (0.8%) in enoxaparin/heparin. The number of reports with the DOAC-levetiracetam-adverse event triplet are listed in table 1.

### Ischemic Stroke

The DOACs did not differ from enoxaparin or heparin in the disproportionality analysis for ischemic stroke, (adj. ROR; 1.14, 95%CI, 1.01-1.28). The adjusted ROR of the interaction effect was 3.57 (95%CI, 2.81–4.58) between DOACs and levetiracetam, supporting the existence of a significant interaction.

When comparing the overall database to each DOAC separately, apixaban and rivaroxaban demonstrated the least disproportionality for ischemic stroke, (adj.ROR; 0.44, 95%CI, 0.41-0.47), and (adj.ROR; 0.35, 95%CI, 0.33-0.37) respectively. A significant disproportionality signal was identified for ischemic stroke in dabigatran (adj. ROR 3.69; 95%CI, 3.50-3.88) and in edoxaban (adj. ROR 1.85; 95%CI, 1.58-2.16) AF users (Table 2).

**Table 2:** Signals of Ischemic Stroke and Anticoagulants-Levetiracetam Drug Interaction in Atrial Fibrillation Patients.

|  | ROR 95%CI | Adj. ROR 95%CI | ROR 95%CI for DDI* |
| --- | --- | --- | --- |
| DOACs vs. Enoxaparin |  |  |  |
| All DOACs | 1.05 (0.94-1.18) | 1.14 (1.01-1.28) | 3.57 (2.81-4.58) |
| DOACs Head-to-Head |  |  |  |
| Apixaban | 0.44 (0.41-0.46) | 0.44 (0.41-0.47) | 4.61 (3.62-5.89) |
| Dabigatran | 3.54 (3.38-3.70) | 3.69 (3.50-3.88) | 4.34 (3.41-5.52) |
| Edoxaban | 1.77 (1.58-1.98) | 1.85 (1.58-2.16) | 5.59 (4.23-7.39) |
| Rivaroxaban | 0.36 (0.35-0.39) | 0.35 (0.33-0.37) | 3.69 (2.92-4.67) |

The first row illustrates the four DOACs compared with enoxaparin. The rows below compared each of the DOACs to the other three head-to-head. \*DDI= drug-drug interaction.

### DOAC-Levetiracetam Drug-Drug Interactions

Apixaban, dabigatran, edoxaban, and rivaroxaban illustrated a significant signal for the drug interaction in the regression model. The strength of association seems higher in edoxaban (adj.ROR 5.59; 95%CI, 4.23-7.39). Whereas the strength of association was slightly lower with rivaroxaban, (adj. ROR 3.69; 95%CI, 2.92-4.67).

When queried for ischemic stroke Ω_0.25_ was greater than zero for apixaban, dabigatran, and rivaroxaban, demonstrating an interaction with levetiracetam (figure 1). The magnitude of Ω_0.25_ was similar for the three DOACs. An interaction between levetiracetam and enoxaparin/heparin was not detected, with Ω_0.25_<0.

**Figure 1:**
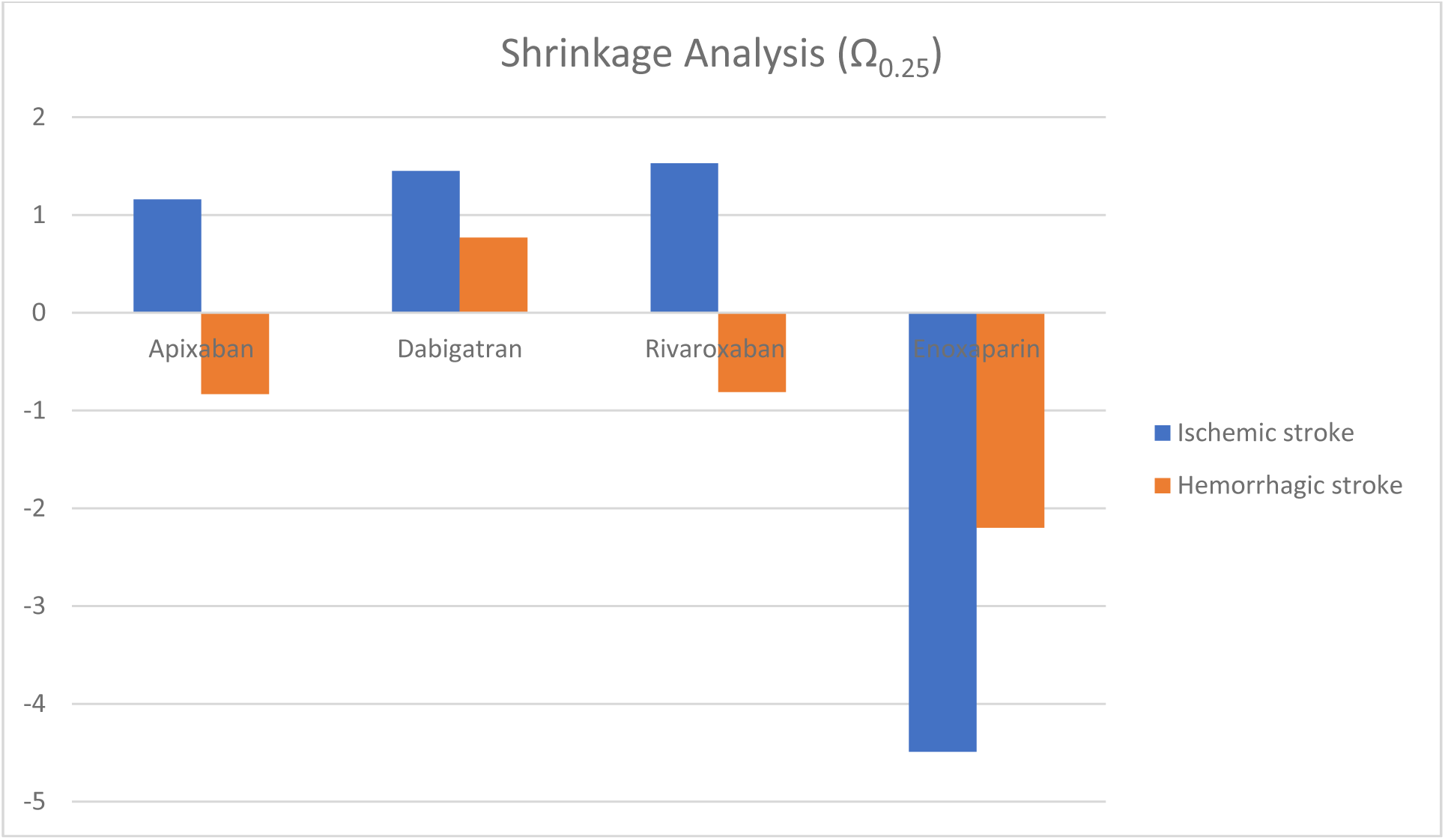
Shrinkage Analysis for the Anticoagulants-Levetiracetam Interaction Ω_025_ > 0, a positive lower bound of 95% CI, is used as a threshold for detecting the signals of the concomitant use of the DOACs and levetiracetam. A positive shrinkage was noticed for ischemic in the concomitant use of either apixaban, dabigatran, and rivaroxaban with levetiracetam. A negative shrinkage was noticed for hemorrhagic stroke and the concomitant use of apixaban or rivaroxaban with levetiracetam.

### Non-Ischemic Stroke- Hemorrhage

A significant disproportionality signal was identified for hemorrhagic stroke in rivaroxaban AF patients compared to apixaban and dabigatran (adj. ROR 1.46; 95%CI, 1.38-1.55) (Table 3). Dabigatran demonstrated a significantly lower adj. ROR for hemorrhagic stroke than the other DOACs, (adj. ROR 0.75; 95%CI, 0.70-0.78). Apixaban did not differ significantly from rivaroxaban and dabigatran.

**Table 3:**
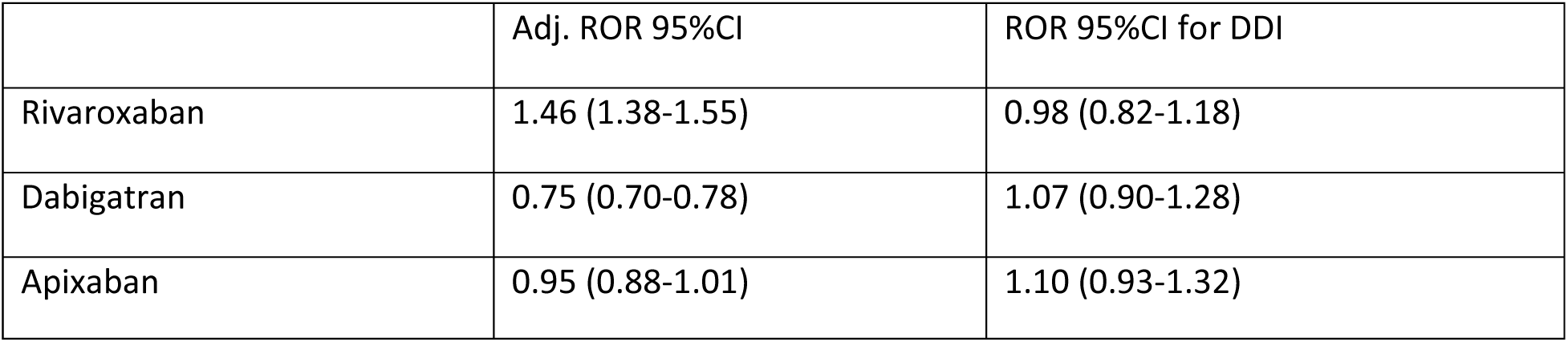
Secondary Outcome Signals of Hemorrhagic Stroke and the DOACs-Levetiracetam Drug Interaction in Atrial Fibrillation Patients.

The regression model queried for hemorrhagic stroke did not detect an interaction with levetiracetam. Likewise, the shrinkage analysis was negative for apixaban, and rivaroxaban, detecting no interaction. The concomitant use of levetiracetam and the two DOACs does not seem to increase the risk of intracerebral bleeding reports. The shrinkage analysis was tested positive for dabigatran (figure 1). Contrary to the shrinkage analysis, the regression model did not detect signals for dabigatran-levetiracetam-hemorrhagic stroke triplet (adj. ROR 1.07; 95%CI, 0.90-1.28).

### Sensitivity Analysis

In sensitivity analysis, we could not detect changes in disproportionality by publication date. The model detected signals in the positive control carbamazepine but not in the negative control, omeprazole (Table 4).

**Table 4:** Sensitivity Analysis for DOAC DDI for Ischemic or non-Ischemic Stroke in Atrial Fibrillation Patients.

| | ROR 95%CI for DDI | Shrinkage Analysis ($\Omega_{0.25}$) | Rationale |
| --- | --- | --- | --- |
| Apixaban-Carbamazepine | 8.47 (5.37-13.36) | +1.97 | Positive control: the concomitant use of apixaban with strong CYP3A4 and P-glycoprotein inducers such as carbamazepine, may lead to a ~50% reduction in apixaban exposure. In a clinical study of atrial fibrillation patients, diminished efficacy was noted (14). The lower bound CI >1 and $\Omega_{0.25}$>0 for ischemic stroke detecting a signal. |
| Apixaban-Omeprazole | 1.13 (0.98-1.33) | -0.45 | Negative control: no interaction is documented to date; hemorrhagic stroke ROR is non-significant. The lower bound CI <1 and $\Omega_{0.25}$<0. |
| Publication date |  |  |  |
| Rivaroxaban-levetiracetam<br>2012-2020 | 3.51 (2.27-5.43) | +2.30 | First, in 2020, Guistozzi et al. illustrated that patients with non-valvular AF treated with rivaroxaban and levetiracetam appear to have a relatively high rate of thromboembolic events (17). Thus, the data was re-analyzed for 2012-2020.<br>Second, a lawsuit occurred over Pradaxa, dabigatran's trade name, from 2010 to 2014 (18, 31). An additional analysis excluding that period did not significantly alter the results. |
| Dabigatran-levetiracetam<br>2015-2023 | 3.14 (2.28-3.42) | +1.31 |  |

## DISCUSSION

The main finding of the current analysis is a significant signal for interaction between levetiracetam and DOACs, as demonstrated by the increased risk of ischemic stroke in the FAERS database. The interaction was confirmed using two methods accepted by regulators worldwide suggesting a robust and significant finding.

The mechanism of the DDI remains to be elucidated. However, it is hypothesized that the mechanism involves P-glycoprotein induction, an efflux protein that lowers the bioavailability of DOACs (8,19). Moreover, levetiracetam may bear a weak enzyme-inducing properties and may stimulate the activity of CYP3A4 and/or some <u>UDP</u>-glucuronosyltransferase (UGT) isoenzymes (19).

Apixaban, dabigatran, edoxaban, and rivaroxaban are all substrates of the efflux P-gp transporter. Our analysis successfully demonstrated an interaction with all four DOACs, with 3-5 folds increased risk of stroke 3-5 and some variability between the DOACs. Rivaroxaban showed the weakest association, probably because of its high baseline bioavailability compared with the other DOACs.

Recently, there has been extensive concern regarding the interaction between levetriacetam and the DOACs. The 2021 European Heart Rhythm Association (EHRA) guide on the use of non-vitamin K antagonist oral anticoagulants in patients with atrial fibrillation recommends caution in the use of the antiepileptic drug, levetiracetam, due to potential P-glycoprotein-mediated drug–drug interaction (8,33). To date, data on the potential drug interaction between levetiracetam and anticoagulants from the DOAC family is scarce.

Paciullo et al. reported a case in 2020 of a 69-year-old man with AF who received rivaroxaban (20 mg/d) and developed a transient ischemic attack a few months after initiating levetiracetam for the indication of focal seizures prevention (20). Specific anti-Xa activity for Rivaroxaban found a zero-trough level immediately before rivaroxaban administration. In 2020, Giustozzi et al. illustrated in a small prospective study (n=91) that patients with non-valvular AF treated with DOACs and anticonvulsants appear to have a relatively high rate of thromboembolic events (17). A similar conclusion was drawn in a nested case-control study supporting a diminished anticoagulant effect when combining DOACs with levetiracetam (21).

On the other hand, recent pharmacokinetic studies suggest there is no such potential drug interaction. In a case study published by Menichelli et al. (22) a 54-year-old male with AF, cirrhosis, and seizures showed no significant reduction in dabigatran plasma concentration when used alongside levetiracetam. Another recent small PK study by Mavri et al. analyzed 21 patients concurrently receiving levetiracetam and DOACs, with 19 having atrial fibrillation and 2 having venous thromboembolism (23). Blood samples were collected to measure trough concentrations of DOACs and levetiracetam. The results showed that none of the patients experienced thromboembolic events during the observation period of 1,388 ± 994 days. Furthermore, there was no reduction in DOAC plasma levels during levetiracetam treatment, suggesting that levetiracetam may not significantly affect DOAC concentrations.

However, this study has two important limitations: first, because DOAC trough levels are highly variable, there is a need to analyze the area under the curve to rule out P-gp induction effect on plasma levels with and without levetiracetam. Second, the reported average trough levels of levetiracetam in that study (31.0 ± 34.5 mg/L) although the average concentration is higher than expected, the standard deviation may not ensure that all DOAC patients were adequately exposed to levetiracetam. In the literature the lower bound confidence interval of levetiracetam concentrations is about 15 mcg/mL (34).

In another retrospective cohort study published by Ip et al., the risk of thromboembolism was evaluated in patients taking direct oral anticoagulants concurrently with antiseizure medications that modulate the cytochrome P450 or P-glycoprotein systems including levetiracetam. While CYP/P-gp-modulating antiseizure medications were associated with an increased risk of ischemic stroke in the overall analysis, no difference in thromboembolism risk or death was observed in the epilepsy subgroup using levetiracetam (24).

Earlier studies on levetiracetam showed no alterations in plasma concentrations of other P-glycoprotein substrates, such as digoxin (25, 26). Thus, it may also be reasonable to investigate pharmacodynamic interaction. Piracetam, a molecule similar in structure to levetiracetam, has been shown to have anticoagulant properties (27). However, our analysis did not show an increase in the reporting ratio of hemorrhagic stroke, defined as major bleeding, not supporting any levetiracetam anticoagulant properties or increased bleeding.

It is worthwhile to note that dabigatran demonstrated a positive shrinkage signal for hemorrhagic stroke with concomitant use of levetiracetam, however the logistic regression failed to support any drug-drug interaction. A retrospective cohort of patients from Taiwan on DOACs and 11 different anticonvulsants reported increased association of bleeding with concomitant prescription of phenytoin, valproic acid or levetiracetam, the results were explained by an increased risk of renal failure with levetiracetam (35). Further, our shrinkage analysis did not detect an interaction with heparin/enoxaparin that resulted in bleeding.

Although an interaction may exist between DOACs and levetiracetam this does not mean physicians should stop prescribing the combination for post stroke epilepsy. Levetiracetam is considered a newer and safer anticonvulsant (36). We recommend a drug monitoring strategy to be implemented as a solution for this drug interaction.

Whether the interaction is pharmacokinetic or pharmacodynamic, also has clinical implications for therapeutic drug monitoring. Naturally, a pharmacokinetic interaction is best monitored by plasma levels of DOACs. Whereas anti-Xa for apixaban and rivaroxaban and the plasma diluted thrombin time for dabigatran would be more suitable for a pharmacodynamic interaction (28, 29).

Regarding monitoring methods, Goldstein et al. examined the impact of cytochrome P450- and P-glycoprotein-inducing antiseizure medications on the pharmacokinetics of direct oral anticoagulants compared to rifampicin (a very strong inducer of DOACs elimination) (30). They suggested monitoring DOAC plasma concentrations as a helpful strategy to guide dosing and identify patients at risk for low DOAC concentrations and treatment failure when taking enzyme-inducing antiseizure medications. In the event our findings are confirmed in further studies and the mechanism for the levetiracetam interaction will be fully understood, pharmacodynamic monitoring should be preferred.

Dabigatran demonstrated a higher adjROR for ischemic stroke than apixaban, edoxaban and rivaroxaban, which could be explained by dabigatran’s negative publicity a year after it accessed the market in 2010-2011 (31). The FAERS database shows a peak with the highest rates of reports between 2011-2014 (18). To account for this, we re-analyzed the data excluding the years 2010-2014, but the signal for ischemic events and the levetiracetam drug interaction did not significantly change. Our results also show that dabigatran has the lowest adj. ROR for hemorrhagic stroke, a reciprocal image of the high reporting on ischemic stroke. In the literature, dabigatran does not seem to differ in effectiveness from the other DOACs but is associated with lower risk of major bleeding (32).

### Strengths and Limitations

The major strengths of our study are utilizing a worldwide database, an analysis using two well validated methods for signal detection, performing an extensive sensitivity analysis to challenge our model and mitigate reporting bias, and finally, suggesting pharmacological mechanism that can explain the findings.

A limitation of our study is that some adverse events are likely not reported to national authorities for inclusion in the FAERS. This was mitigated by collecting data from all countries from 2012 to 2023. Although concomitant drug use was accounted for in our analysis, it is challenging to determine with certainty the sequence of medication use owing to missing data and temporality being associated with an adverse event rather than a drug. Another caveat is that reports lack information on dose.

Although lamotrigine is an alternative medication to post-stroke epilepsy, its use for that indication is seemingly low. There were no ischemic stroke reports in DOACs combined with lamotrigine, therefore we used omeprazole as our negative control, which is usually given to prevent gastrointestinal bleeding with DOACs. The analysis could not detect a drug interaction with DOACs and omeprazole, strengthening our results.

There were no reports of intracranial bleeding with the concomitant use of levetiracetam and edoxaban, limiting the direction analysis for edoxaban-levetiracetam interaction. Although, the regression model and shrinkage analysis detected an interaction with edoxaban, a discrepancy in their magnitudes were noted. This could be explained by a high proportion of ischemic stroke reports and a low number of non-cases reporting concomitant levetiracetam use, which emphasized this variable in the regression model.

Finally, limitations to spontaneous reporting (e.g., underreporting) exist as well. However, despite its flaws, the analysis of pharmacovigilance databases remains a cornerstone for the study of adverse drug reactions and drug interactions by regulators and contributes majorly to drug labels. Data science in medicine is growing fast and is especially sensitive for detecting variables that may interfere with drug therapy.

### Conclusions and Clinical Implications

We show a strong signal for the levetiracetam interaction with apixaban, dabigatran, edoxaban, and rivaroxaban. The interaction is demonstrated by 3-5 folds increased reporting risk of ischemic stroke. Our findings suggest the need for pharmacodynamic monitoring, either anti-Xa for apixaban and rivaroxaban or plasma diluted thrombin time for dabigatran while concomitantly prescribing levetiracetam with the DOACs.

## Data Availability Statement

*The data underlying this article are available in* the FDA adverse event reporting system (FAERS) at https://www.fda.gov/drugs/questions-and-answers-fdas-adverse-event-reporting-system-faers/fda-adverse-event-reporting-system-faers-public-dashboard.

## Notes

Funding/Support: This research received no specific grant from any funding agency in the public, commercial, or not-for-profit sectors.

### Competing Interest Statement

The authors have declared no competing interest.

### Funding Statement

No funding was provided for this project.

### Author Declarations

No oversight body since it is an open public database FAERS analysis.

